# Metabolomic signatures of data-driven type 2 diabetes subtypes and their associations with dementia and stroke risk

**DOI:** 10.64898/2026.08.21.26361082

**Authors:** Si Han, Jack Hewett, Geert Jan Biessels, Fariba Ahmadizar

## Abstract

**Background:** Data-driven type 2 diabetes (T2D) subtypes differ in their risks of dementia and stroke. We examined whether their metabolomic profiles also differed and whether subtype-related metabolic patterns were associated with dementia, stroke, and all-cause mortality.

**Methods:** We analyzed NMR-based metabolomic profiles across previously defined T2D subtypes in the UK Biobank. Subtype-related metabolites were summarized using principal component analysis (PCA), and their associations with incident dementia, stroke, and all-cause mortality were examined using Cox models. Attenuation analyses and two-sample Mendelian randomization further assessed subtype-outcome relationships and the potential causal relevance of outcome-associated metabolites.

**Results:** Among 7,671 individuals (mean age 59.85 years; 37% female), the first five PCs explained 76.7% of variance in subtype-related metabolites and mainly reflected lipid and lipoprotein signatures. After adjustment for T2D subtype and confounders, the HDL-remodeling PC increased risks of all-cause dementia (HR 1.17, 95% CI 1.08-1.27), VaD (HR 1.18, 95% CI 1.05-1.32), and all-cause mortality (HR 1.16, 95% CI 1.13-1.19). Lower scores on the LDL cholesterol-enriched axis increase risks of all-cause dementia (HR 0.75, 95% CI 0.62-0.91) and mortality (HR 0.76, 95% CI 0.69-0.83). The VLDL/LDL-enriched PC was inversely associated with mortality (HR 0.93, 95% CI 0.88-0.98). No significant stroke results were observed. Adjustment for the PCA-derived metabolomic patterns generally attenuated subtype-outcome associations, MR analyses identified 197 metabolite-outcome associations that remained significant after FDR correction.

**Conclusions:** Metabolomic profiling showed that the metabolic signatures differed across data-driven T2D subtypes and highlighted lipid and lipoprotein remodeling as a major metabolic feature associated with dementia, stroke, and all-cause mortality.

**Research in context:** *What is this study about?:* This study examined metabolomic signatures across previously defined data-driven type 2 diabetes (T2D) subtypes and their associations with dementia, stroke, and all-cause mortality. Using NMR-based metabolomics in the UK Biobank, we identified distinct metabolic patterns that differed across T2D subtypes and were related to long-term outcomes.

*What is novel about this study?:* This study integrates comprehensive NMR metabolomics with data-driven T2D subtypes to identify metabolic patterns associated with dementia, stroke, and mortality. Rather than focusing on individual metabolites, we identified biologically interpretable metabolomic patterns and showed that differences in lipid and lipoprotein composition were prominent features of subtype heterogeneity and long-term outcome risk.

*What is the main take-home message?:* Data-driven T2D subtypes exhibit distinct metabolomic profiles, largely driven by lipid and lipoprotein metabolism. Metabolomic signatures reflecting high-density lipoprotein (HDL) remodeling and lipoprotein composition were consistently associated with dementia, stroke, and all-cause mortality.

*What is the clinical implication?:* Metabolomic profiling may complement conventional T2D classification by improving biological characterization of metabolic heterogeneity and identifying candidate biomarkers for future risk stratification and precision prevention.

**Highlights:**

- NMR metabolomics distinguished data-driven T2D subtypes in the UK Biobank.
- Subtype differences were largely driven by lipid and lipoprotein remodeling.
- HDL patterns were associated with dementia, stroke, and all-cause mortality.
- Metabolomics may improve T2D phenotyping and future risk stratification.

## Introduction

Type 2 diabetes (T2D) is a clinically and biologically heterogeneous disease, primarily defined by abnormal glucose levels, that arises from the interaction of metabolic, vascular, and inflammatory pathways (1, 2). Yet, current diagnostic and treatment approaches remain largely uniform and do not fully account for individual pathophysiological differences (3–5). This heterogeneity likely contributes to the variability in disease progression and clinical outcomes.

Data-driven clustering has identified reproducible T2D subgroups with distinct metabolic features, including obesity- and inflammation-related, vascular aging, and mild metabolic phenotypes (6–8), routine clinical features are also used for individualized prediction models (9). In our previous study, we identified severe obesity-related inflammatory diabetes (SOID), mild metabolic diabetes (MMD), and mild age-related hypertension-predominant diabetes (MARD-H) (10). These subtypes differed in their subsequent risks of stroke and dementia, indicating clinically relevant variation in long-term neurovascular outcomes (10). Although these subtypes highlight substantial heterogeneity within T2D, also in terms of risk of stroke and dementia, it remains unclear whether distinct metabolomic signatures characterize these subtypes and whether such signatures are associated with differences in long-term neurovascular outcomes.

Metabolomics captures systemic biochemical variation related to lipid metabolism, amino acid turnover, oxidative stress, and energy balance (11, 12). Linking metabolomic profiles to clinically derived T2D subtypes may therefore help clarify the biological basis of subtype heterogeneity and its relationships with adverse outcomes.

In this study, we characterized the metabolomic profiles of T2D subtypes previously established through unsupervised clustering in a large cohort of individuals with T2D in the UK Biobank (UKB) using comprehensive nuclear magnetic resonance (NMR)-based metabolomics. We aimed to (1) derive major subtype-related metabolic patterns using principal component analysis and compare them across T2D subtypes; (2) examine their associations with dementia, stroke, and all-cause mortality; (3) assess attenuation of subtype–outcome associations after metabolomic adjustment; and (4) assess the potential causal relevance of subtype-related metabolites associated with dementia, stroke, and all-cause mortality using Mendelian Randomization (MR).

## Methods

### 2.1. Study setting and study population

The UKB is a population-based prospective cohort of more than 500,000 participants aged 40 to 69 years at recruitment between 2006 and 2010. The study received ethics approval from the Northwest Multicenter Research Ethics Committee (reference no. 16/NW/0274), and all participants provided written informed consent. At 22 assessment centers across England, Scotland, and Wales, participants completed questionnaires and interviews, underwent physical measurements, and provided biological samples.

Participants with diabetes at baseline were identified by integrating multiple data sources, as described in previous studies (13, 14) (*Supplementary Figure S1*). Hospital data were obtained from Hospital Episode Statistics for England and Wales and Scottish Morbidity Records for Scotland. T2D was identified using ICD-9 codes 250.00, 250.10, 250.20, and 250.90 or ICD-10 code E11. The baseline assessment date was used as the index date, and demographic characteristics, covariates, medication use, and metabolomic measurements were derived from this visit.

### 2.2. Metabolomics profiling

Detailed protocols for sample collection and metabolomic quantification have been described previously (15, 16). Plasma metabolites were measured in UKB baseline EDTA-plasma aliquot 3 samples using the Nightingale Health high-throughput NMR platform. Analyses were conducted in three phases between June 2019 and November 2023 using nine NMR spectrometers operated by Nightingale Health (Helsinki, Finland).

Data underwent centralized quality control and preprocessing before release. Biomarker values were analyzed as provided by UKB without additional batch correction, in accordance with platform recommendations. Biomarkers with more than 20% missing values were excluded; the remaining biomarkers were natural log-transformed and standardized as z-scores. Baseline blood samples were collected under non-fasting conditions at the same assessment as the clinical variables used for clustering. Although aliquot 3 samples may have undergone approximately 5-10% dilution during handling, this is unlikely to materially affect epidemiological association analyses (17).

### 2.3. Definition of T2D subtypes

T2D subtypes were defined previously, using six routinely measured clinical variables: age at diagnosis, body mass index (BMI), HbA1c, insulin resistance (IR) estimated using the triglyceride-to-HDL cholesterol ratio, systolic blood pressure (SBP), and C-reactive protein (CRP) (9). Missing values were addressed using random forest imputation. Right-skewed variables (CRP and IR) were log-transformed to reduce skewness. All six variables were then standardized to z-scores to ensure equal weighting in the clustering process. Unsupervised k-means clustering was performed, and the optimal number of clusters was determined via the elbow method based on within-cluster sum of squares (WSS). To assess structural robustness, hierarchical clustering using Ward’s D^2^ linkage was applied in parallel. The details can be found in our previous study (10).

### 2.4 Outcomes

Incident dementia and stroke outcomes were identified using algorithmically defined classifications provided by UKB, including incident dementia (all-cause, Alzheimer’s disease [AD], vascular dementia [VaD]) and stroke (all-cause, ischemic [IS], and hemorrhagic [ICH]), defined using hospital, primary care, and death records. These classifications were based on a combination of self-reported medical conditions at baseline (Field 20002), hospital inpatient records coded using ICD-10 (Data-Coding 2000), and death register data (Fields 40001 and 40002). The algorithms were developed by the UKB outcome adjudication group to ensure high positive predictive value, enabling consistent and reliable identification of health-related events across multiple data sources (18). Death data were obtained from the death certificates from the National Health Service Information Center in England and Wales and the National Health Service Central Register in Scotland. Follow-up time was calculated from the date of baseline assessment to the date of the outcome’s diagnosis, death, or the censoring date, whichever occurred first. To minimize potential reverse causation, events occurring within the first two years following the baseline assessment date were excluded.

### 2.5 Covariates

Covariates were selected a priori based on established associations with cognitive, vascular, and metabolic health. These included demographic factors (age at baseline, sex, and ethnicity), socioeconomic position (Townsend deprivation index and educational attainment), lifestyle factors (smoking status, alcohol consumption and an overall diet quality score reflecting habitual dietary intake), use of lipid-lowering, blood pressure-lowering, glucose-lowering medications, and kidney function, assessed by estimated glomerular filtration rate (eGFR) calculated using the CKD-EPI 2021 equation, to evaluate the robustness of associations to potential confounding by treatment use and diabetes-related comorbidity.

### 2.6 Statistical analysis

#### 2.6.1 Baseline characteristics

Baseline characteristics were summarized according to T2D subtype. Continuous variables are presented as means (standard deviations) and medians (interquartile ranges), whereas categorical variables are presented as frequencies and percentages.

#### 2.6.2 Identification of subtype-related metabolites

Associations between individual metabolite concentrations and T2D subtype were examined using multivariable linear regression models. Metabolite concentrations were log-transformed where appropriate and standardized to a mean of 0 and a standard deviation of 1 before analysis. T2D subtype was included as a categorical exposure variable.

For each metabolite, two nested multivariable linear regression models were fitted. The reduced model included the covariates only, whereas the full model additionally included T2D subtypes. The overall contribution of T2D subtype was evaluated using a partial F-test comparing the full and reduced models. This test assessed the null hypothesis that the covariate-adjusted mean metabolite concentration was equal across all three T2D subtypes. A significant overall test therefore indicated that at least one subtype differed from another. Metabolites with an overall FDR-adjusted q value <0.05 were classified as subtype-related metabolites and were selected for the subsequent principal component analysis.

For the selected metabolites, pairwise contrasts were estimated between SOID and MMD, MARD-H and MMD, and SOID and MARD-H to characterize the direction and magnitude of the subtype differences. MMD was used as the reference subtype for model estimation. Pairwise P values were corrected using the Benjamini-Hochberg procedure separately within each contrast and adjustment model. Model 1 was adjusted for age, sex, ethnicity, educational attainment, deprivation index, smoking status, alcohol consumption, and dietary factors. Model 2 additionally included kidney function and medication use.

#### 2.6.3 PCA-derived subtype-related metabolic patterns

Principal component analysis (PCA) was applied to summarize the correlated structure of these subtype-related metabolites into a smaller number of coordinated metabolomic patterns (19, 20), using selected metabolites from the previous section 2.6.2. Metabolite concentrations were log-transformed where appropriate and standardized to a mean of 0 and a standard deviation of 1, so that subsequent effect estimates represented a 1-standard-deviation increase in the component score. Because the proportion of missing metabolite data was low, PCA was performed using complete cases. Principal components (PCs) were retained based on inspection of the scree plot and the eigenvalue criterion. Component scores were calculated for each participant, and PCs were interpreted according to the metabolites with the largest absolute loadings.

The distributions of the retained PCA-derived component scores were summarized across the three T2D subtypes. Adjusted differences in component scores were examined using multivariable linear regression, with MMD as the reference subtype and the same covariate adjustment in previous analyses.

#### 2.6.4 PCA-derived metabolic patterns and outcomes

Associations between the retained PCA-derived metabolomic patterns and incident dementia, stroke, and all-cause mortality were examined using Cox proportional hazards regression models. Each standardized principal component score was entered separately as a continuous exposure, and hazard ratios (HRs) and 95% confidence intervals (CIs) were estimated per 1-standard-deviation increase in the component score. Same confounder-models were applied with further adjusted for T2D subtype, to determine whether the metabolomic patterns were associated with clinical outcomes independently of subtype classification. The proportional hazards assumption was assessed using Schoenfeld residuals.

#### 2.6.5 Attenuation of subtype-outcome associations after adjustment for metabolomic patterns

To assess whether PCA-derived metabolomic patterns accounted for subtype differences in dementia, stroke, and mortality risk, subtype-outcome associations were estimated before and after adjustment for all retained PCs. For each outcome, Cox models with identical samples were fitted with prespecified covariates, first without and then with the PC scores. Percentage attenuation was calculated from the change in subtype coefficients on the log-hazard scale as:

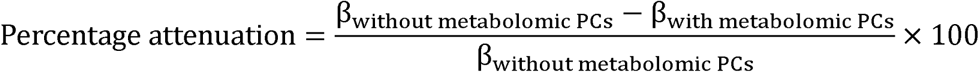

A positive attenuation value indicated that the subtype coefficient moved toward the null after adjustment for the metabolomic patterns, whereas a negative value indicated strengthening of the association. Attenuation estimates were not emphasized when the original subtype coefficient was close to zero or when the direction of the association changed after adjustment. These estimates were interpreted as the proportion of the observed subtype-outcome association statistically accounted for by the PCA-derived metabolomic patterns, rather than as evidence of formal causal mediation.

#### 2.6.6 Mendelian Randomization analyses

Two-sample MR was performed for subtype-related metabolites associated with dementia, stroke, and all-cause mortality. Genetic instruments were obtained from published metabolite genome-wide association studies (GWAS) (21, 22) and included independent genome-wide significant variants (P < 5 × 10^−8^; r² < 0.01 within 10,000 kb). Instrument strength was assessed using F-statistics. Outcome summary statistics were obtained from relevant GWAS datasets (21–25), and exposure and outcome alleles were harmonized. The primary analysis used inverse-variance weighting (IVW) with multiplicative random effects. Sensitivity analyses were performed using MR-Egger, weighted-median, and weighted-mode methods where sufficient instruments were available. Heterogeneity and horizontal pleiotropy were assessed using Cochran’s Q and the MR-Egger intercept test, respectively.

#### 2.6.7 Sensitivity analysis

Several sensitivity analyses were conducted to assess the robustness of the findings. First, associations between individual metabolite concentrations (z-scores) and outcomes were also evaluated using Cox proportional hazards models adjusted for T2D subtype and the same covariates as in the primary outcome analyses. Second, to reduce the potential influence of reverse causation, all outcome analyses were repeated after excluding events occurring within the first five years following the baseline assessment. Third, to minimize potential circularity related to the lipid-derived insulin resistance proxy used in subtype derivation, we excluded metabolites highly correlated with HDL-C, triglycerides, or the insulin resistance proxy. Spearman correlation coefficients were calculated between each metabolite and these three variables. Metabolites with an absolute correlation coefficient greater than 0.80 with any of these variables were excluded, and the metabolite–outcome analyses were repeated. PCA was then repeated using the remaining metabolites, and the resulting metabolomic patterns and their associations with T2D subtypes and clinical outcomes were re-evaluated.

All analyses were performed using R (version 4.3) with the cmprsk, cluster, and TwoSampleMR packages. Multiple testing was addressed using the Benjamini-Hochberg FDR method. Statistical significance was evaluated using FDR-adjusted thresholds (q < 0.05 and q < 0.10).

## Results

### 3.1 Cluster features

In the present analysis, 7,671 participants with available plasma metabolomics data were included. Three previously identified T2D subtypes were fitted: severe obesity-related inflammatory diabetes (SOID; n=2,397), mild metabolic diabetes (MMD; n=2,807), and mild age-related hypertension-predominant diabetes (MARD-H; n=2,467) (10). Baseline cluster characteristics of the three subtypes are shown in *Table 1*. The full characteristics can be found in *Supplementary Table S1*.

**Table 1.** Clinical characteristics of the data-driven T2D subtypes in the original and metabolomics cohorts. (a) Original cohort (n = 14,353)

| Variable | **SOID**<br>Severe obesity-related<br>inflammatory T2D<br>(n = 4,515) | **MMD**<br>Mild metabolic<br>T2D<br>(n = 5,148) | **MARD-H**<br>Mild age-related<br>hypertension-predominant T2D<br>(n = 4,690) |
| --- | --- | --- | --- |
| Age at T2D diagnosis (years, mean (SD)) | 53.84 (7.65) | 54.99 (7.61) | 61.60 (5.13) |
| BMI (kg/m<sup>2</sup>, mean (SD)) | 36.56 (6.07) | 28.83 (4.22) | 30.61 (4.32) |
| HbA1c (mmol/mol, mean (SD)) | 58.2 (13.9) | 49.1 (9.2) | 51.3 (8.6) |
| Triglyceride-to-HDL cholesterol ratio (proxy for insulin resistance), mean (SD)) | 2.54 (1.24) | 1.35 (0.78) | 1.85 (0.92) |
| SBP (mmHg, mean (SD)) | 137.89 (14.64) | 130.49 (11.91) | 156.37 (14.42) |
| CRP (mg/L, mean (SD)) | 4.74 (4.27) | 1.17 (1.11) | 1.91 (1.75) |

(b) Metabolomics cohort (n = 7,671)
| Variable | **SOID**<br>Severe obesity-related<br>inflammatory T2D<br>(n = 2,397) | **MMD**<br>Mild metabolic<br>T2D<br>(n = 2,807) | **MARD-H**<br>Mild age-related<br>hypertension-predominant T2D<br>(n = 2,467) |
| --- | --- | --- | --- |
| Age at T2D (years, mean (SD)) | 53.89 (7.72) | 55.06 (7.72) | 61.65 (5.10) |
| BMI (kg/m<sup>2</sup>, mean (SD)) | 36.57 (6.00) | 28.90 (4.22) | 30.60 (4.30) |
| HbA1c (mmol/mol, mean (SD)) | 61.65 (18.18) | 49.25 (10.36) | 52.63 (11.61) |
| Insulin resistance (TG/HDL ratio, unitless, mean (SD)) | 3.08 (2.09) | 1.44 (0.85) | 2.08 (1.40) |
| SBP (mmHg, mean (SD)) | 137.90 (14.40) | 130.50 (11.92) | 156.38 (14.28) |
| CRP (mg/L, mean (SD)) | 7.40 (8.24) | 1.63 (1.77) | 2.69 (3.17) |

Age at T2D diagnosis was calculated as the difference between the recorded diagnosis date and the participant's date of birth. All continuous variables were standardized (z-scored) before clustering to ensure equal weighting across features; however, values are displayed here on the original measurement scale for interpretability. Statistical comparisons are descriptive only, as these variables directly contributed to the clustering process and differences between clusters are therefore expected by design. No inferential statistical testing was performed because cluster-defining variables were used to derive the clusters.
Abbreviations: T2D, type 2 diabetes; BMI, body mass index; HbA1c, glycated hemoglobin; TG/HDL, triglyceride-to-high-density lipoprotein cholesterol ratio (proxy for insulin resistance); SBP, systolic blood pressure; CRP, C-reactive protein.

### 3.2 Identification of subtype-related metabolites

A total of 251 circulating metabolites and lipid-related measures were evaluated for differences across the MMD, SOID, and MARD-H subtypes. In the fully adjusted model, overall subtype differences were observed for 243 of the 251 metabolites after F-test, corresponding to 96.8% of all evaluated measures (all q<0.05) (*Supplementary Table S2*).

The strongest overall subtype associations were observed for the ratio of polyunsaturated to monounsaturated fatty acids (F=742.11, q=1.08*10^−291^), the percentage of monounsaturated fatty acids relative to total fatty acids (F=688.22, q=2.00*10^−272^), the percentage of polyunsaturated fatty acids relative to total fatty acids (F=682.27, q=1.98*10^−270^), and glycoprotein acetyls (F=671.95, q=8.74*10^−267^). Strong differences were also found for several HDL-, LDL-, triglyceride-, and fatty-acid composition measures.

Pairwise analyses demonstrated extensive metabolic separation among the three subtypes (*Supplementary Table S3*): 230 of 243 subtype-related metabolites differed between SOID and MMD, 219 between MARD-H and MMD, and 201 between SOID and MARD-H. Of these, 167 were significant in all three comparisons, 73 in two, and 3 in one.

Compared with MMD, SOID had lower polyunsaturated-to-monounsaturated fatty-acid ratio (β=-0.97, 95% CI [−1.02, −0.92]) and polyunsaturated fatty-acid percentage (β=-0.95, 95% CI [−1.00, −0.90]), but higher glycoprotein acetyls (β=0.95, 95% CI [0.90, 1.00]), monounsaturated fatty-acid percentage (β=0.93, 95% CI [0.88, 0.98]), and triglycerides as a percentage of total lipids in small HDL (β=0.92, 95% CI [0.87, 0.97]). MARD-H showed higher triglyceride-rich and VLDL-related measures than MMD, with the largest differences around β=0.48-0.49. Compared with MARD-H, SOID had higher medium-HDL phospholipid percentage, glycoprotein acetyls, and large-LDL triglyceride percentage, but lower polyunsaturated-to-monounsaturated, polyunsaturated, and omega-6 fatty-acid percentages.

### 3.3 PCA-derived subtype-related metabolic patterns

Principal component analysis was performed using the 243 metabolites that showed a significant overall association with diabetes subtype in the fully adjusted model. Parallel analysis supported retention of 18 principal components, whereas 21 components had eigenvalues greater than 1 (*Supplementary Table S4*). Parallel analysis was used as the primary retention criterion. PC1 and PC2 explained 32.3% and 23.7% of the total variance, respectively, and together accounted for 56.0% of the variation in the subtype-related metabolite profile. PC3, PC4, and PC5 explained a further 10.7%, 6.2%, and 3.8%, respectively. Thus, the first five components jointly explained 76.7% of the total variance and captured the major correlated metabolic patterns underlying the subtype-related metabolite differences.

PC1 (*Figure 1-a*) was dominated by the abundance and lipid content of triglyceride-rich lipoproteins. Strong loadings included total VLDL particle concentration and lipid measures across small, medium, and large VLDL subclasses, together with VLDL cholesterol, LDL triglycerides, and circulating fatty acids. PC1 therefore represented a broad VLDL/LDL-enriched pattern, with higher scores indicating greater VLDL particle and lipid concentrations and higher triglyceride- and fatty acid-related measures (*Supplementary Table S5*).

**Figure 1.**
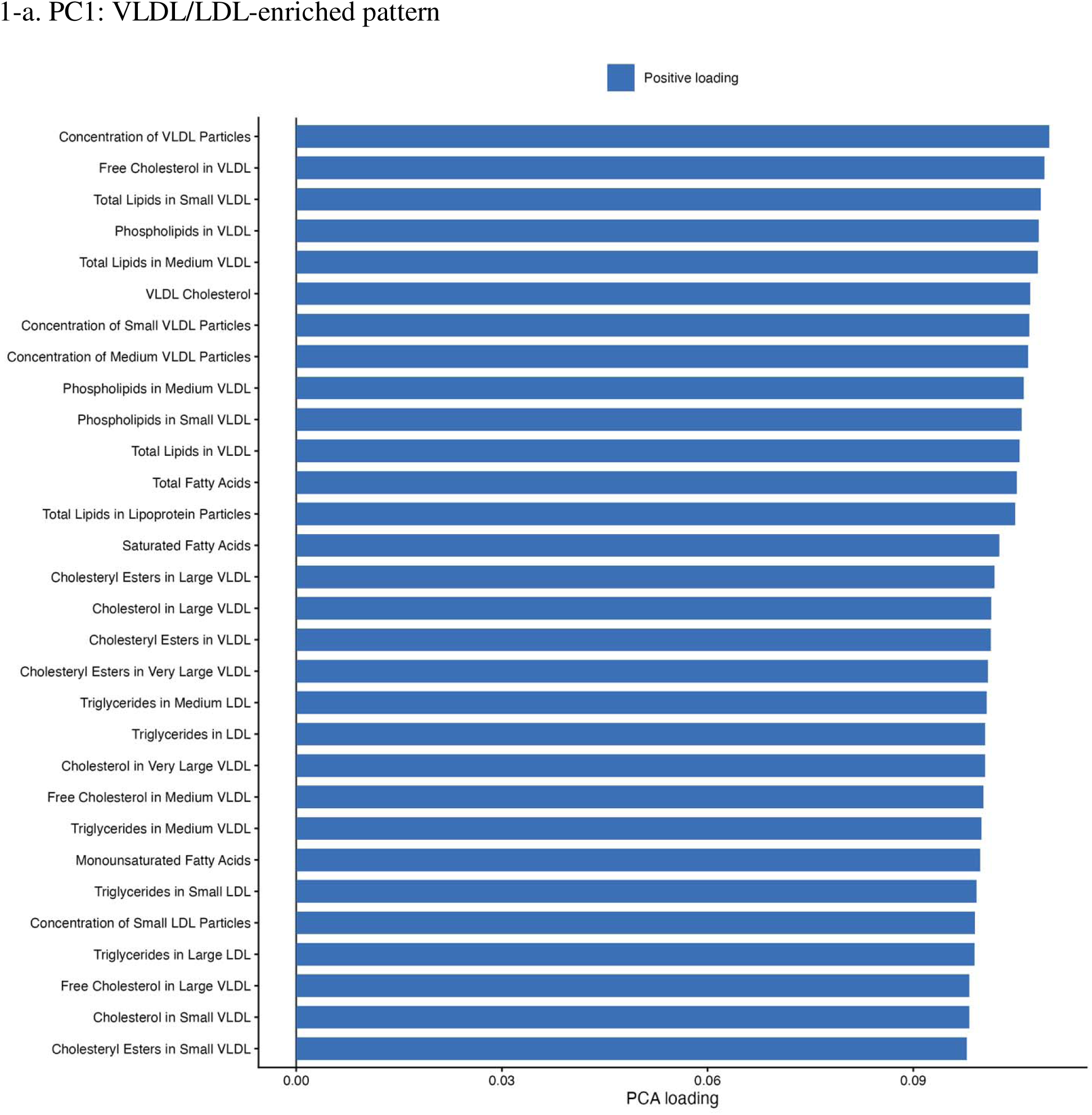
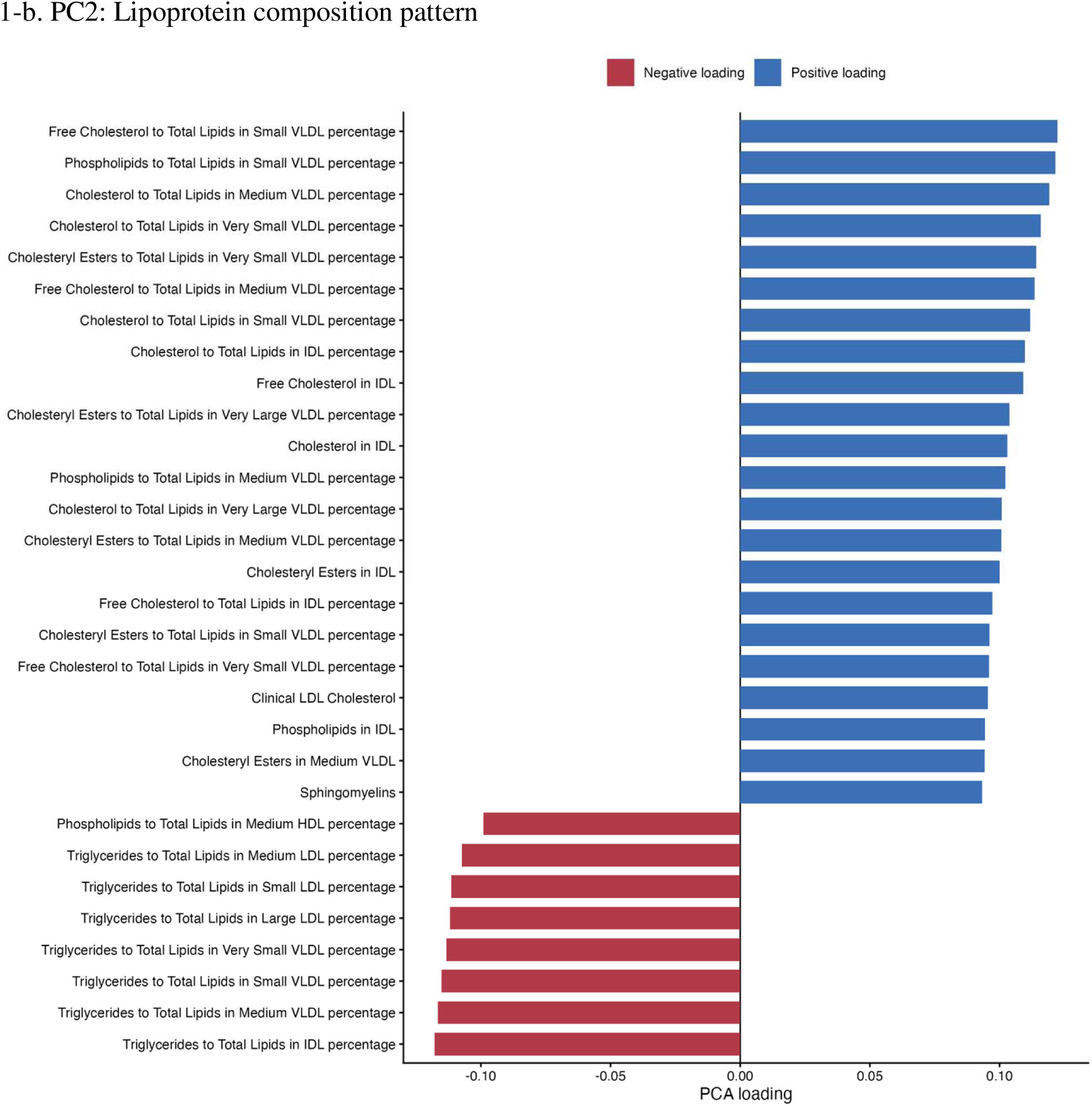
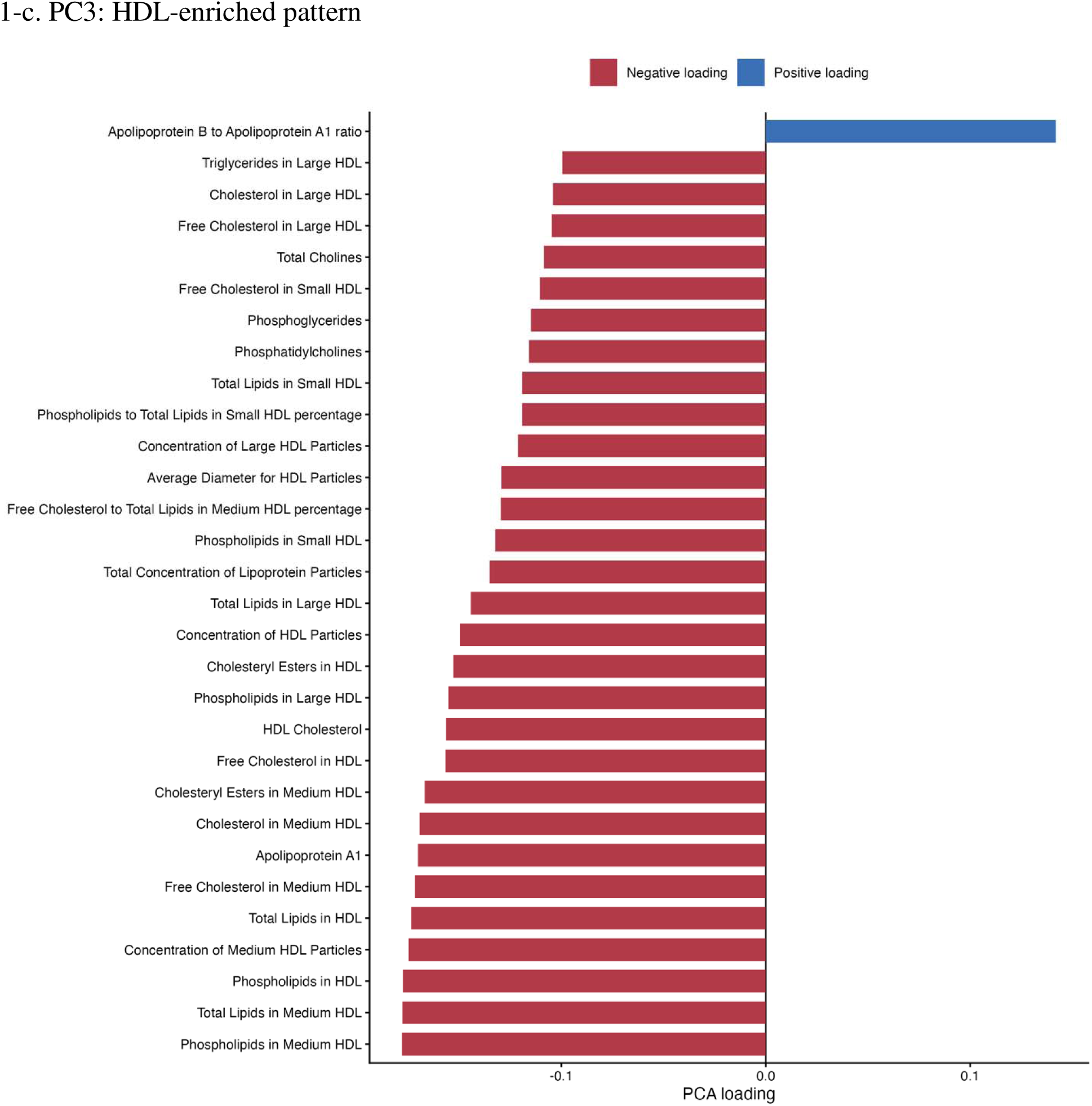
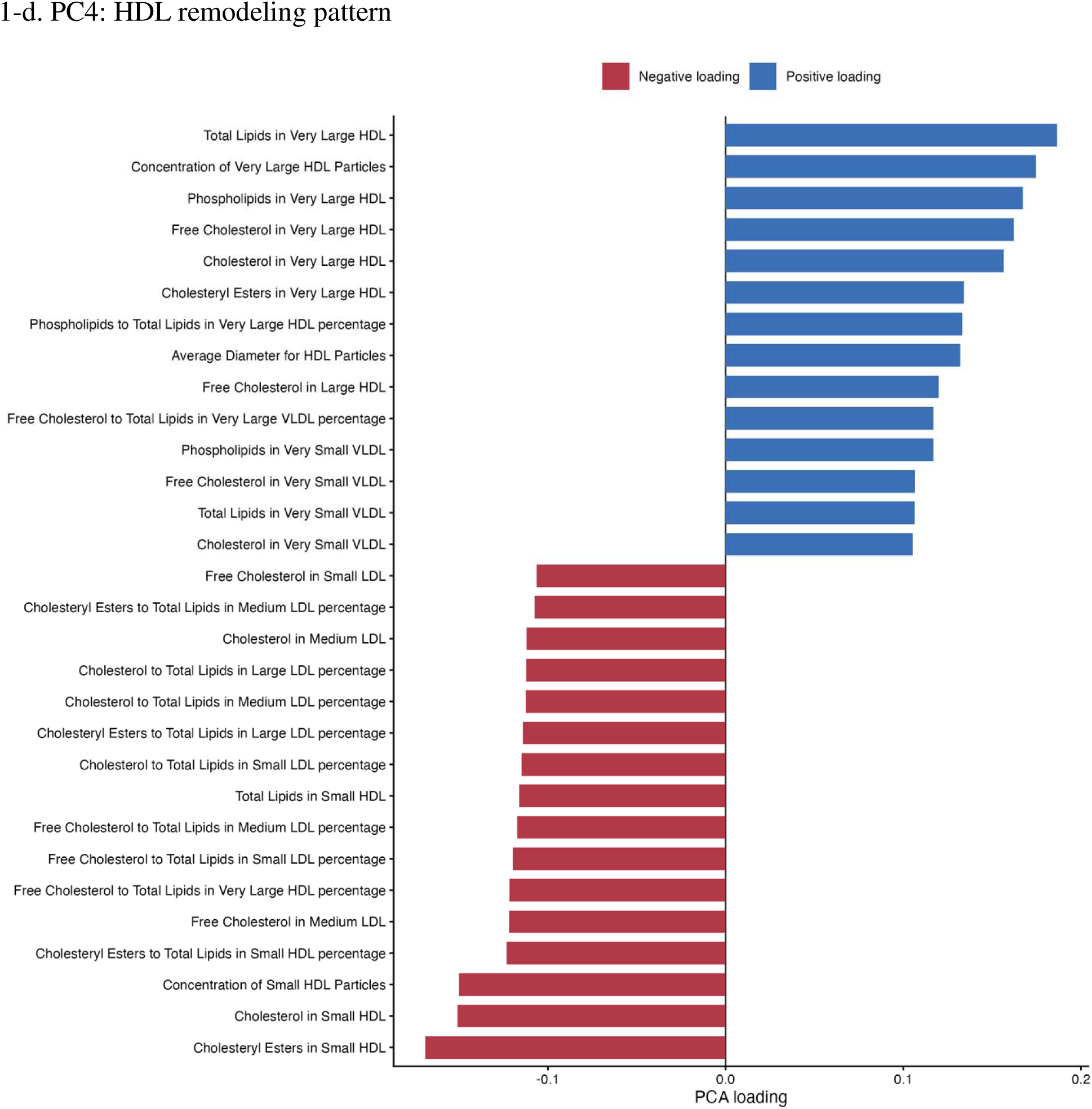
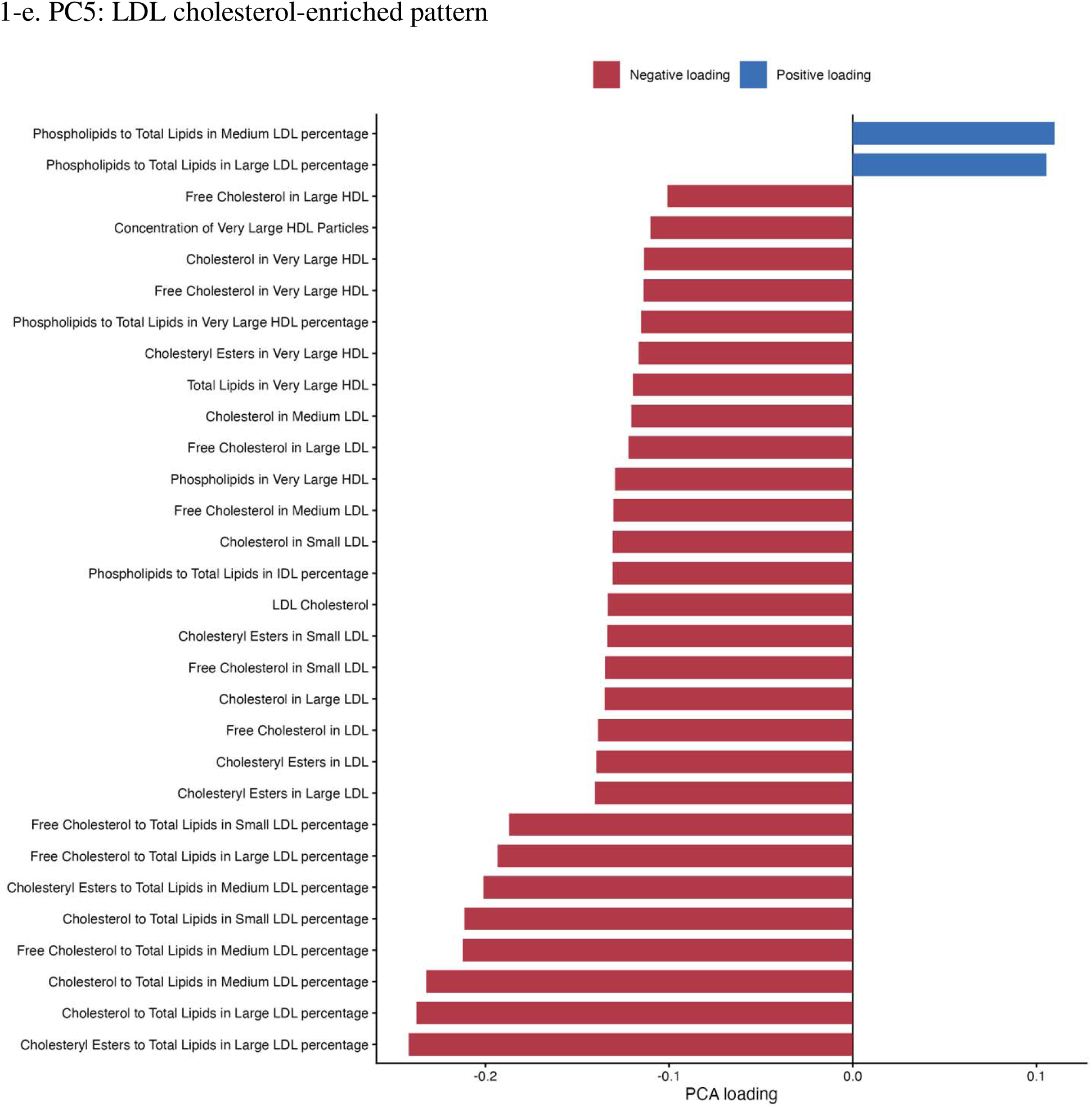
Standardized PCA- derived metabolomic signatures in participants with type 2 diabetes The top metabolites with the largest absolute loadings are shown for each of the first five principal components (PCs), which together explained 76.7% of the total variance in circulating metabolites. Bars represent the metabolites with the largest absolute loadings. Positive and negative loadings indicate metabolites contributing in opposite directions to each principal component. Abbreviations: PCA, principal component analysis; PC, principal component; VLDL, very-low-density lipoprotein; IDL, intermediate-density lipoprotein; LDL, low-density lipoprotein; HDL, high-density lipoprotein; TG, triglycerides; ApoA1, apolipoprotein A1; ApoB, apolipoprotein B

PC2 (*Figure 1-b* mainly reflected relative lipoprotein composition rather than absolute concentration. Cholesterol, free cholesterol, and phospholipid proportions in VLDL and IDL loaded positively, whereas triglyceride proportions in VLDL, IDL, and LDL loaded negatively. Higher PC2 scores therefore indicated cholesterol-, free cholesterol-, and phospholipid-enriched particles with lower relative triglyceride content, while lower scores reflected a more triglyceride-enriched composition (*Supplementary Table S5*).

PC3 (*Figure 1-c*) was predominantly HDL-related, with strong contributions from HDL lipids, HDL particle concentrations, HDL size, HDL cholesterol, and apolipoprotein A1. The apolipoprotein B-to-apolipoprotein A1 ratio loaded in the opposite direction. PC3 was interpreted as an HDL-enriched pattern. In the current orientation, lower scores indicated higher HDL particle concentrations, lipid content, particle size, and apolipoprotein A1, whereas higher scores reflected the opposing profile. The sign of the component is arbitrary (*Supplementary Table S5*).

PC4 (*Figure 1-d*) contrasted very large and small HDL particles. Very large HDL particle concentration, lipid content, cholesterol, free cholesterol, phospholipids, and HDL particle diameter loaded positively, whereas small HDL particle and lipid measures loaded negatively. PC4 was therefore interpreted as an HDL remodeling pattern, with higher scores indicating a shift toward larger HDL particles and lower scores indicating relative predominance of small HDL particles (*Supplementary Table S5*).

PC5 (*Figure 1-e*) reflected LDL cholesterol composition. Cholesterol, free cholesterol, and cholesteryl esters in large, medium, and small LDL loaded strongly, while phospholipid proportions in LDL loaded in the opposite direction. PC5 was interpreted as an LDL cholesterol-composition pattern. In the current orientation, lower scores indicated greater LDL enrichment with cholesterol, free cholesterol, and cholesteryl esters, whereas higher scores indicated relatively greater phospholipid content (*Supplementary Table S5*).

### 3.4 PCA-derived metabolic patterns and outcomes

In models additionally adjusted for diabetes subtype, several PCA-derived metabolic patterns were associated with incident dementia subtypes and all-cause mortality after correction for multiple testing (*Figure 2*) (*Supplementary Table S6*).

**Figure 2.**
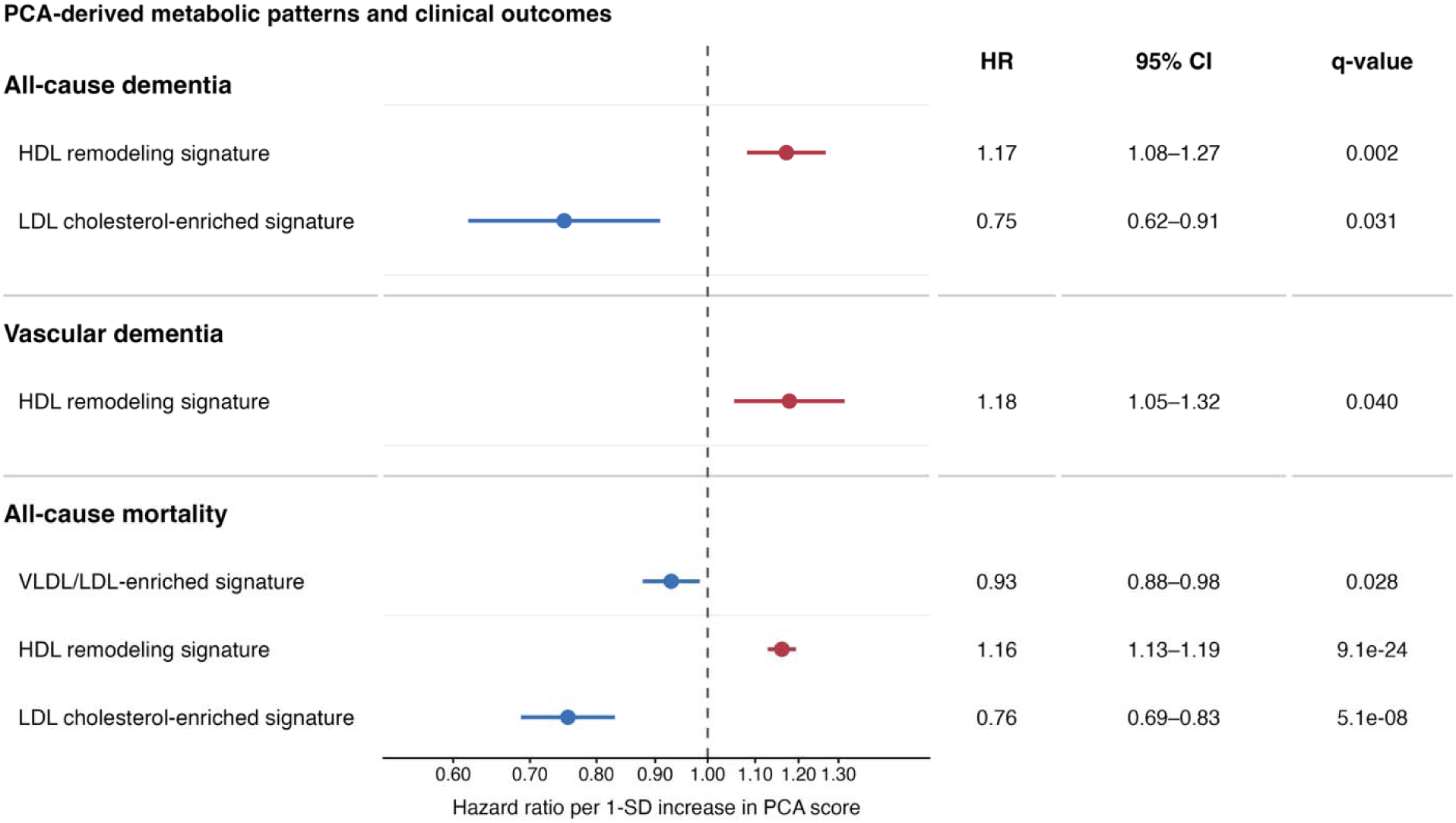
Significant associations of PCA-derived metabolomic patterns with clinical outcomes Points represent hazard ratios per 1-SD increase in principal component (PC) score, and horizontal bars indicate 95% confidence intervals. Only associations remaining significant after false discovery rate (FDR) correction are shown. Hazard ratios were estimated using fully adjusted Cox proportional hazards models (model 2). Abbreviations: PCA, principal component analysis; PC, principal component; VLDL, very-low-density lipoprotein; IDL, intermediate-density lipoprotein; LDL, low-density lipoprotein; HDL, high-density lipoprotein

For all-cause dementia, higher HDL remodeling signature scores were associated with an increased risk, with an HR of 1.17 per SD increase in the PC score (95% CI 1.08-1.27; q=0.0016). In contrast, higher LDL cholesterol-enriched signature scores were associated with a lower risk of all-cause dementia (HR 0.75, 95% CI 0.62-0.91; q=0.031). Given the loading structure of these components, the HDL remodeling signature association indicated that a metabolic profile characterized by a relative predominance of very large HDL particles, larger HDL particle size, and lower levels of small HDL-related measures was associated with higher all-cause dementia risk. The inverse association for LDL cholesterol-enriched signature indicated that lower scores, corresponding to greater LDL cholesterol, free cholesterol, and cholesteryl ester enrichment, were associated with higher all-cause dementia risk. For VaD, higher HDL remodeling signature scores were similarly associated with increased risk (HR 1.18, 95% CI 1.05-1.32; q=0.040), showed consistent positive associations with both all-cause dementia and VaD.

The largest number of significant associations was observed for all-cause mortality. Higher HDL remodeling signature scores were strongly associated with increased mortality risk (HR 1.16, 95% CI 1.13-1.19; q=9.07*10^−24^). In contrast, higher scores for VLDL/LDL-enriched signature were associated with lower mortality risk. Specifically, the HRs were 0.93 for PC1 (95% CI 0.88-0.98; q=0.028). The direction of the LDL cholesterol-enriched signature association was consistent across all-cause dementia and mortality. Because higher LDL cholesterol-enriched signature scores represented relatively lower LDL cholesterol and cholesteryl ester enrichment, the inverse HRs indicated that the opposing LDL cholesterol-enriched signature profile, characterized by greater LDL cholesterol enrichment was associated with higher risks of both all-cause dementia and mortality. By contrast, higher HDL remodeling signature scores, reflecting a shift toward very large HDL particles and away from small HDL particles, were consistently associated with increased risks of all-cause dementia, VaD, and mortality. No significant associations were observed in stroke outcomes.

### 3.5 Attenuation of subtype-outcome associations after adjustment for metabolomic patterns

Adjustment for the PCA-derived metabolomic patterns generally attenuated subtype-outcome associations, although the extent of attenuation differed substantially between subtype contrasts and outcomes (*Figure 3*) (*Supplementary Table S6*).

**Figure 3.**
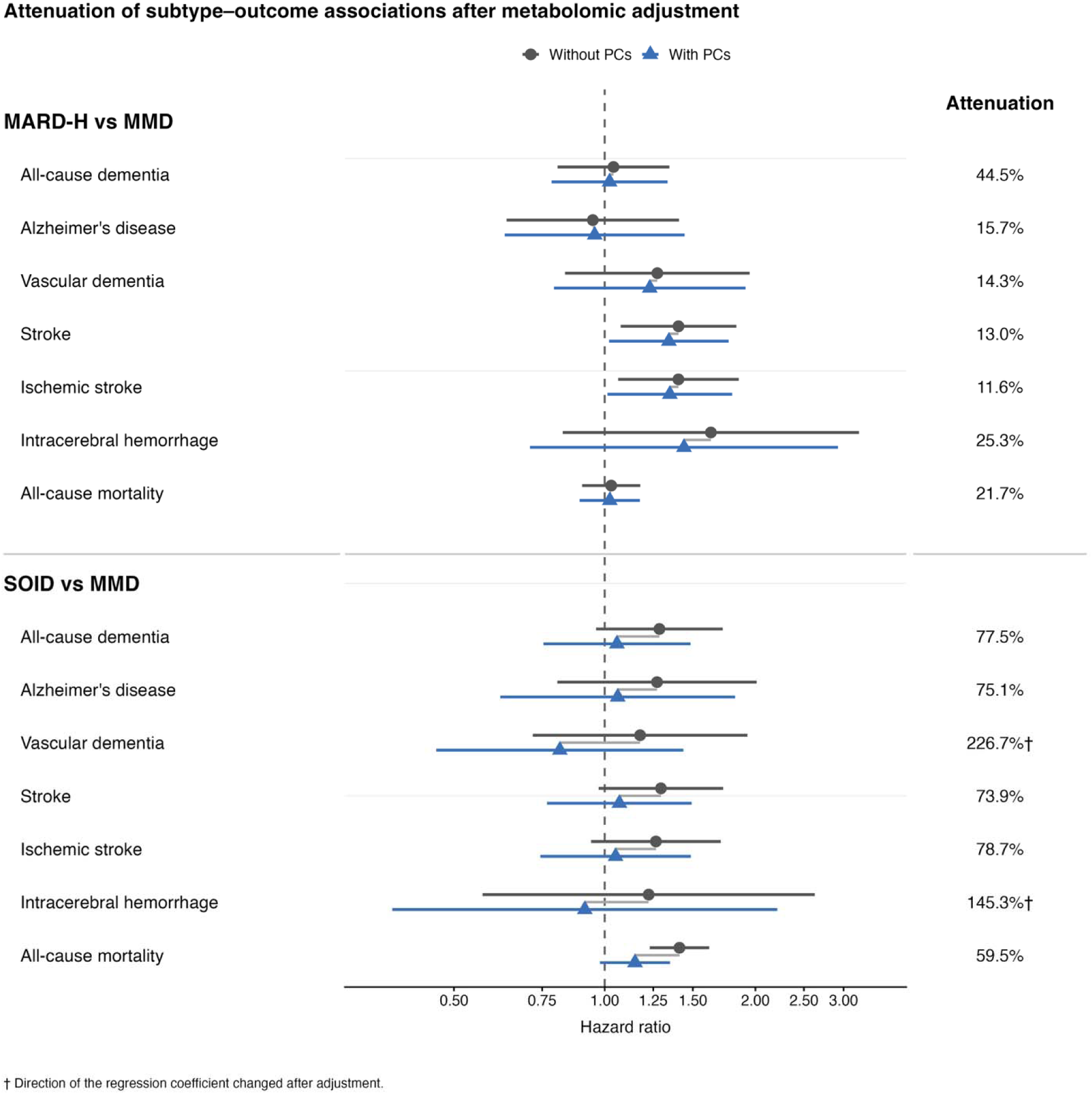
Attenuation of subtype-outcome associations after adjustment for PCA-derived metabolomic patterns Hazard ratios and 95% confidence intervals are shown for associations of MARD-H and SOID, each compared with MMD, before and after additional adjustment for PCA-derived metabolomic patterns. Points represent hazard ratios and horizontal lines represent 95% confidence intervals. Connecting lines indicate the change in effect estimates after metabolomic adjustment. Attenuation was calculated on the log-hazard scale. Values greater than 100% indicate that the regression coefficient crossed the null and changed direction after adjustment.

For SOID versus MMD, most associations were markedly attenuated after adjustment for metabolomic patterns. Attenuation was 77.5% for all-cause dementia, from HR 1.29 (95% CI 0.96-1.72) to 1.06 (0.75-1.48); 75.1% for AD, from 1.27 (0.80-2.01) to 1.06 (0.62-1.82); 78.7% for IS, from 1.27 (0.94-1.71) to 1.05 (0.74-1.49); and 73.9% for stroke, from 1.30 (0.97-1.72) to 1.07 (0.77-1.49). Mortality was attenuated by 59.5%, from HR 1.41 (1.23-1.62; p=8.05*10^−7^) to 1.15 (0.98-1.35; p=0.089), and was no longer statistically significant. For ICH and VaD, estimates crossed the null after adjustment, but confidence intervals were wide and associations were non-significant both before and after adjustment.

For MARD-H versus MMD, attenuation was smaller: 44.5% for all-cause dementia, 15.7% for AD, 25.3% for ICH, 21.7% for mortality, and 14.3% for VaD. Associations with IS and stroke remained significant after adjustment. For IS, the HR decreased from 1.40 (95% CI 1.06-1.85; p=0.017) to 1.35 (1.01-1.80; p=0.041), corresponding to 11.6% attenuation. For stroke, the HR decreased from 1.40 (1.08-1.83; p=0.012) to 1.34 (1.02-1.77; p=0.035), corresponding to 13.0% attenuation.

### 3.6 Mendelian Randomization analysis

Primary IVW MR analyses identified 197 metabolite-outcome associations that remained significant after FDR correction. These comprised 79 associations with AD, 48 with all-cause dementia, 47 with all-cause mortality, and 23 with stroke. The number of genetic instruments contributing to individual estimates ranged from 8 to 110 SNPs (*Supplementary Table S7*).

For all-cause dementia, 48 estimates remained FDR-significant, including 38 positive and 10 inverse associations. The strongest positive association was for the cholesterol proportion in large LDL (OR 1.44, 95% CI 1.24-1.68; q<0.001). Higher LDL cholesterol, total lipids in LDL, and free cholesterol in medium LDL were also associated with greater risk. Inverse associations were observed for the phospholipid proportion in IDL and triglyceride proportions in medium LDL, IDL, large LDL, and small LDL. For AD, 79 estimates were FDR-significant, including 68 positive and 11 inverse associations. Strong positive associations were observed for lipid measures in small and medium LDL, including phospholipids, free cholesterol, total lipids, cholesterol, and cholesteryl esters (ORs 3.66-4.22; all q<0.001), as well as LDL cholesterol (OR 3.73, 95% CI 2.07-6.75; q<0.001). Inverse associations were observed for the phospholipid proportion in small HDL, triglyceride proportions in VLDL, LDL, and IDL subclasses, and glutamine.

For stroke, 21 of 23 FDR-significant associations were positive. Higher apolipoprotein B to apolipoprotein A1 ratio, free cholesterol in medium LDL, LDL cholesterol, cholesterol in small LDL, and clinical LDL cholesterol were associated with higher risk (ORs 1.10-1.11; q=0.045). Cholesteryl esters and cholesterol in medium HDL were inversely associated with stroke.

For all-cause mortality, all 47 FDR-significant estimates indicated higher risk and mainly involved cholesterol-rich VLDL, IDL, and LDL traits. The strongest evidence was observed for phospholipids in IDL, cholesterol and cholesteryl esters in medium VLDL, total free cholesterol, and the free cholesterol proportion in small HDL (ORs 1.11-1.17; q=0.006). Higher apolipoprotein B, LDL particle concentration, clinical LDL cholesterol, total cholesterol, non-HDL cholesterol, remnant cholesterol, and multiple LDL lipid measures were also associated with increased mortality risk.

### 3.7 Sensitivity analysis

In the individual metabolite analyses, several metabolites were associated with dementia, stroke, and all-cause mortality *(Supplementary Table S8)*. All-cause dementia risk was higher with very large HDL measures but lower with triglycerides in very large VLDL and small HDL measures. A higher free cholesterol-to-total lipids ratio in small HDL was associated with stroke (HR 1.21, 95% CI 1.10-1.33). For mortality, glucose, pyruvate, 3-hydroxybutyrate, and very large HDL particles were positively associated, whereas LDL cholesterol, triglyceride- and fatty acid-related measures, branched-chain amino acids, and small HDL measures were inversely associated.

In sensitivity analyses applying a 5-year lag, the direction and magnitude of associations were broadly consistent with the primary findings, although fewer associations remained significant. Lipoprotein-related metabolites continued to show the most consistent associations with dementia and all-cause mortality *(Supplementary Table S9)*.

To reduce potential circularity from the lipid-based insulin resistance proxy used for subtype derivation, PCA was repeated after excluding 80 metabolites highly correlated with HDL-C, triglycerides, or the insulin resistance proxy (|Spearman *r*| > 0.80). Among the 171 retained metabolites, the first five sensitivity PCs explained 70.1% of total variance and remained predominantly lipid-related *(Supplementary Table S10)*. T2D subtype differences were largely preserved. Compared with MMD, sPC2 scores were higher in SOID (β = 4.45, q < 0.001) and MARD-H (β = 2.30, q < 0.001), while MARD-H had lower sPC1 scores (β = −1.10, q < 0.001). Higher sPC3 and sPC5 scores were also associated with increased all-cause dementia risk (HR 1.03, 95% CI 1.02-1.05; and HR 1.11, 95% CI 1.05-1.18, respectively) *(Supplementary Table S11-12)*.

## Discussion

Our study identified distinct metabolomic signatures across data-driven T2D subtypes, with the most pronounced differences involving lipid and lipoprotein metabolism. Among 251 measured metabolites, 243 were associated with subtype in the overall adjusted analysis, and most also differed in at least two pairwise subtype comparisons. PCA of these subtype-related metabolites identified several biologically interpretable patterns, including VLDL/LDL enrichment, lipoprotein composition, HDL enrichment, HDL remodeling, and LDL cholesterol enrichment. These findings indicate that the clinical subtype framework captures broad and coordinated metabolic differences rather than variation in a small number of isolated biomarkers.

The clearest metabolic separation was observed for SOID compared with MMD. SOID showed widespread differences in triglyceride-rich lipoproteins, VLDL-related measures, LDL-related measures, and lipoprotein composition. This profile is compatible with the greater obesity and insulin resistance that characterize SOID, because insulin resistance may promote hepatic VLDL production, impair triglyceride-rich lipoprotein clearance, and alter lipid exchange among lipoprotein subclasses (26, 27). MARD-H also differed from MMD across a large number of metabolites, although its metabolic profile appeared less pronounced than that of SOID. The large number of metabolites shared across pairwise contrasts suggests that the subtypes differ along several overlapping metabolic axes rather than through entirely subtype-specific pathways.

The PCA results further demonstrated that subtype-related metabolic variation was clinically relevant beyond subtype classification itself. After adjustment for cluster membership and conventional risk factors, the HDL-remodeling pattern was associated with higher risks of all-cause dementia, VaD, and all-cause mortality. The LDL cholesterol-enriched pattern was associated with all-cause dementia and mortality, while the VLDL/LDL-enriched pattern was associated with mortality. Because the direction of a PC is arbitrary, these associations should be interpreted according to the loading structure rather than the numerical sign of the component alone. In the orientation used in this study, higher HDL remodeling signature scores reflected greater contributions from very-large and larger HDL measures and lower contributions from small-HDL measures. Its positive associations with dementia and mortality therefore indicate that variation in HDL subclass distribution and remodeling, rather than simply higher or lower total HDL cholesterol, was related to adverse outcomes. The association of the LDL cholesterol-enriched pattern with dementia and mortality also requires interpretation in relation to its loading orientation. In the current PCA solution, LDL cholesterol, free cholesterol, and cholesteryl ester measures contributed predominantly to the negative side of the LDL cholesterol-enriched signature. Consequently, the inverse HRs for LDL cholesterol-enriched signature indicate that the LDL cholesterol-enriched side of this metabolic axis was associated with higher outcome risk.

Adjustment for PCA-derived patterns attenuated SOID associations with dementia and stroke by approximately 74%-79% and mortality by 59.5%, after which the mortality association was no longer significant. This suggests that metabolomic patterns captured part of the excess risk associated with SOID, although the analysis does not establish mediation. MR findings also implicated lipid and lipoprotein metabolism. Higher genetically associated LDL-, IDL-, and VLDL-related traits were associated with dementia, stroke, and mortality, with the strongest associations for AD. However, correlated NMR traits, pleiotropy, exposure scaling, and overlapping instruments limit causal interpretation and warrant replication.

Our findings are relevant to the continuing debate regarding HDL and vascular risk. The observational results did not support a simple interpretation in which higher HDL-related measures are uniformly protective. Instead, associations depended on HDL subclass, lipid composition, and the metabolic axis represented by the PCA score. The HDL-remodeling component distinguished very-large and larger HDL measures from small-HDL measures and was positively associated with dementia and mortality. Similarly, the MR results showed heterogeneous associations across HDL-related traits rather than a consistent protective effect of all HDL measures. This pattern is compatible with the concept that HDL particle composition and remodeling may provide information not captured by total HDL cholesterol alone (28–32). Nevertheless, the present analyses assess circulating traits and genetically proxied metabolite levels rather than direct measures of HDL function, and conclusions regarding cholesterol efflux, anti-inflammatory capacity, or other functional properties cannot be drawn from these data.

The mortality findings were dominated by lipid and lipoprotein measures. Although the broader metabolomic dataset included non-lipid metabolites, the results provided here do not support describing inflammation, glycolysis, ketone-body metabolism, or renal dysfunction as the principal mortality-associated patterns. The most consistent evidence instead involved cholesterol-rich LDL, IDL, and VLDL traits in the MR analyses and the VLDL/LDL-enriched, HDL-remodeling, and LDL cholesterol-enriched PCA patterns in the observational analyses. The discussion should therefore emphasize lipid transport, cholesterol distribution, and lipoprotein remodeling rather than broader multisystem metabolic dysfunction unless additional metabolite-level results demonstrate those pathways.

This study has several strengths, including the use of a large, well-characterized T2D cohort, comprehensive NMR metabolomic profiling, and the integration of data-driven subtype classification with metabolite-level, PCA, attenuation, and MR analyses. Restricting the PCA to metabolites associated with subtype allowed the dimensionality-reduction analysis to focus specifically on metabolic variation relevant to T2D heterogeneity. The use of standardized PC scores facilitated comparison across components, and adjustment for subtype membership enabled evaluation of whether metabolomic patterns contained prognostic information beyond the clinical clusters.

Several limitations should also be considered. Residual confounding and reverse causation remain possible in the observational analyses. Although outcome events occurring shortly after diabetes assessment were excluded in the primary analyses, the results of the additional five-year lag analysis should only be discussed after those estimates have been examined. The attenuation analyses do not establish mediation. PCA components are data-dependent, their signs are arbitrary, and each component represents a combination of correlated metabolites rather than a single pathway. The NMR platform is weighted toward lipid and lipoprotein traits and may underrepresent other biologically relevant processes. The predominantly European-ancestry population may limit generalizability. Finally, the MR analyses were constrained by the available exposure and outcome GWASs and by the high correlation among lipoprotein traits.

These findings have potential implications for the biological characterization of T2D heterogeneity. Conventional subtype definitions summarize differences in age, adiposity, glycemia, insulin resistance, and related clinical features, but the present results demonstrate that substantial metabolic variation remains within and across these categories. Metabolomic profiling may therefore provide complementary information by identifying the lipid-transport and lipoprotein-remodeling processes associated with future dementia, vascular disease, and mortality. The substantial attenuation of SOID-associated risks after adjustment for the PCA patterns further suggests that metabolomic information may help explain why individuals assigned to the same clinically defined subtype experience different long-term outcomes. Clinical implementation, however, will require external validation, assessment of incremental predictive performance, and evaluation of whether metabolomic information changes preventive or therapeutic decisions.

In conclusion, data-driven T2D subtypes were characterized by extensive differences in lipid and lipoprotein metabolism. PCA-derived patterns reflecting HDL remodeling, LDL cholesterol enrichment, and VLDL/LDL-related variation were associated with dementia and all-cause mortality independently of subtype membership and conventional risk factors. These patterns accounted for a substantial proportion of several adverse outcome associations in SOID but explained relatively little of the persistent stroke risk in MARD-H. MR analyses provided complementary evidence implicating genetically proxied LDL-, IDL-, VLDL-, and selected HDL-composition traits in dementia, stroke, and mortality. Collectively, the findings identify lipid distribution and lipoprotein remodeling as central features of metabolic heterogeneity in T2D while underscoring the need for cautious causal interpretation and external validation.

## Author Contributions

S.H. contributed to data curation, formal analysis, investigation, methodology, software, visualization, and writing of the original draft. J.H. contributed to methodology and writing-review and editing. G.J.B. contributed to investigation, methodology, supervision, and writing-review and editing. F.A. contributed to conceptualization, investigation, methodology, project administration, resources, supervision, and writing-review and editing. All authors provided final approval of the version to be published.

## Availability of data and materials

This research was conducted using the UK Biobank resource under application number 100993. The UK Biobank data are available to bona fide researchers upon application and approval by UK Biobank (www.ukbiobank.ac.uk). Restrictions apply to the availability of these data, which were used under license for the current study and are not publicly available.

## Declaration of Generative AI and AI-assisted technologies in the writing process

During the preparation of this work, the authors used ChatGPT (OpenAI) to assist with language editing, improving clarity, and grammar of the text. The authors critically reviewed, edited, and verified all AI-assisted content and take full responsibility for the accuracy, originality, and integrity of the final manuscript.

## Funding

S.H. is supported by a scholarship from the China Scholarship Council (CSC) under Grant No. 202208330062. The study funder was not involved in the study design, the collection, analysis, and interpretation of data, or writing of the report, and did not impose any restrictions regarding the publication of the report.

## Conflicts of Interest

The authors declare that they have no competing interests.

## Notes

### Competing Interest Statement

The authors have declared no competing interest.

### Author Declarations

This research was conducted using the UK Biobank resource under application number 100993. UK Biobank has approval from the North West - Haydock Research Ethics Committee (REC reference: 16/NW/0274).

## References

1. Lu X, Xie Q, Pan X, Zhang R, Zhang X, Peng G, et al. Type 2 diabetes mellitus in adults: pathogenesis, prevention and therapy. Signal Transduct Target Ther. 2024;9(1):262.

2. Galicia-Garcia U, Benito-Vicente A, Jebari S, Larrea-Sebal A, Siddiqi H, Uribe KB, et al. Pathophysiology of Type 2 Diabetes Mellitus. Int J Mol Sci. 2020;21(17).

3. Han S, Naderi E, Wang K, Ma Y, Biessels GJ, Ahmadizar F. Prediabetes as a critical stage for risk of dementia and stroke: evidence from the UK Biobank and Mendelian Randomization. European Journal of Preventive Cardiology. 2026.

4. van Sloten TT, Sedaghat S, Carnethon MR, Launer LJ, Stehouwer CDA. Cerebral microvascular complications of type 2 diabetes: stroke, cognitive dysfunction, and depression. Lancet Diabetes Endocrinol. 2020;8(4):325–36.

5. Leslie RD, Ma RCW, Franks PW, Nadeau KJ, Pearson ER, Redondo MJ. Understanding diabetes heterogeneity: key steps towards precision medicine in diabetes. Lancet Diabetes Endocrinol. 2023;11(11):848–60.

6. Wang J, Liu JJ, Gurung RL, Liu S, Lee J, M Y, et al. Clinical variable-based cluster analysis identifies novel subgroups with a distinct genetic signature, lipidomic pattern and cardio-renal risks in Asian patients with recent-onset type 2 diabetes. Diabetologia. 2022;65(12):2146–56.

7. Misra S, Wagner R, Ozkan B, Schön M, Sevilla-Gonzalez M, Prystupa K, et al. Precision subclassification of type 2 diabetes: a systematic review. Commun Med (Lond). 2023;3(1):138.

8. Ahlqvist E, Storm P, Käräjämäki A, Martinell M, Dorkhan M, Carlsson A, et al. Novel subgroups of adult-onset diabetes and their association with outcomes: a data-driven cluster analysis of six variables. Lancet Diabetes Endocrinol. 2018;6(5):361–9.

9. Mori T, Herder C, Cardoso P, Dennis JM, Kuß O. Type 2 diabetes subtypes for precision medicine: methodological challenges and alternative prediction-based approaches. Diabetologia. 2026.

10. Han S, Zhou Y, Sturkenboom M, Biessels GJ, Ahmadizar F. Data-driven subtypes of type 2 diabetes and risk of dementia, stroke, and brain structural changes in the UK Biobank. Diabetes Res Clin Pract. 2026;237:113311.

11. Rakusanova S, Cajka T. Metabolomics and Lipidomics for Studying Metabolic Syndrome: Insights into Cardiovascular Diseases, Type 1 & 2 Diabetes, and Metabolic Dysfunction-Associated Steatotic Liver Disease. Physiol Res. 2024;73(S1):S165–s83.

12. Newgard CB. Metabolomics and Metabolic Diseases: Where Do We Stand? Cell Metab. 2017;25(1):43–56.

13. Han H, Cao Y, Feng C, Zheng Y, Dhana K, Zhu S, et al. Association of a Healthy Lifestyle With All-Cause and Cause-Specific Mortality Among Individuals With Type 2 Diabetes: A Prospective Study in UK Biobank. Diabetes Care. 2022;45(2):319–29.

14. Eastwood SV, Mathur R, Atkinson M, Brophy S, Sudlow C, Flaig R, et al. Algorithms for the Capture and Adjudication of Prevalent and Incident Diabetes in UK Biobank. PLoS One. 2016;11(9):e0162388.

15. Soininen P, Kangas AJ, Würtz P, Suna T, Ala-Korpela M. Quantitative serum nuclear magnetic resonance metabolomics in cardiovascular epidemiology and genetics. Circ Cardiovasc Genet. 2015;8(1):192–206.

16. Soininen P, Kangas AJ, Würtz P, Tukiainen T, Tynkkynen T, Laatikainen R, et al. High-throughput serum NMR metabonomics for cost-effective holistic studies on systemic metabolism. Analyst. 2009;134(9):1781–5.

17. Nightingale Health Plc. Nightingale Health Metabolic Biomarkers Companion Document for UK Biobank NMR Metabolomics Data (Resource 130). UK Biobank Showcase; 2023. Available from: https://biobank.ndph.ox.ac.uk/ukb/refer.cgi?id=130 [

18. UK Biobank. Algorithmically defined outcomes (ADOs) 2022 [Available from: https://biobank.ndph.ox.ac.uk/showcase/showcase/docs/alg_outcome_main.pdf.

19. Worley B, Powers R. Multivariate Analysis in Metabolomics. Curr Metabolomics. 2013;1(1):92–107.

20. Ringnér M. What is principal component analysis? Nat Biotechnol. 2008;26(3):303–4.

21. Borges MC, Haycock PC, Zheng J, Hemani G, Holmes MV, Davey Smith G, et al. Role of circulating polyunsaturated fatty acids on cardiovascular diseases risk: analysis using Mendelian randomization and fatty acid genetic association data from over 114,000 UK Biobank participants. BMC Med. 2022;20(1):210.

22. Borges MC, Schmidt AF, Jefferis B, Wannamethee SG, Lawlor DA, Kivimaki M, et al. Circulating Fatty Acids and Risk of Coronary Heart Disease and Stroke: Individual Participant Data Meta-Analysis in Up to 16 126 Participants. J Am Heart Assoc. 2020;9(5):e013131.

23. Malik R, Chauhan G, Traylor M, Sargurupremraj M, Okada Y, Mishra A, et al. Multiancestry genome-wide association study of 520,000 subjects identifies 32 loci associated with stroke and stroke subtypes. Nat Genet. 2018;50(4):524–37.

24. Kunkle BW, Grenier-Boley B, Sims R, Bis JC, Damotte V, Naj AC, et al. Genetic meta-analysis of diagnosed Alzheimer’s disease identifies new risk loci and implicates Aβ, tau, immunity and lipid processing. Nat Genet. 2019;51(3):414–30.

25. Kurki MI, Karjalainen J, Palta P, Sipilä TP, Kristiansson K, Donner KM, et al. FinnGen provides genetic insights from a well-phenotyped isolated population. Nature. 2023;613(7944):508–18.

26. Bays HE, Kirkpatrick C, Maki KC, Toth PP, Morgan RT, Tondt J, et al. Obesity, dyslipidemia, and cardiovascular disease: A joint expert review from the Obesity Medicine Association and the National Lipid Association 2024. Obes Pillars. 2024;10:100108.

27. Feingold KR. Obesity and Dyslipidemia. [Updated 2026 Mar 24]. In: Feingold KR, Adler RA, Ahmed SF, et al., editors. Endotext [Internet]. South Dartmouth (MA): MDText.com, Inc.; 2000-. Available from: https://www.ncbi.nlm.nih.gov/books/NBK305895/.

28. Hottman DA, Chernick D, Cheng S, Wang Z, Li L. HDL and cognition in neurodegenerative disorders. Neurobiol Dis. 2014;72 Pt A:22–36.

29. Stukas S, Robert J, Wellington CL. High-density lipoproteins and cerebrovascular integrity in Alzheimer’s disease. Cell Metab. 2014;19(4):574–91.

30. Voight BF, Peloso GM, Orho-Melander M, Frikke-Schmidt R, Barbalic M, Jensen MK, et al. Plasma HDL cholesterol and risk of myocardial infarction: a mendelian randomisation study. Lancet. 2012;380(9841):572–80.

31. Rohatgi A, Khera A, Berry JD, Givens EG, Ayers CR, Wedin KE, et al. HDL cholesterol efflux capacity and incident cardiovascular events. N Engl J Med. 2014;371(25):2383–93.

32. Rohatgi A, Westerterp M, von Eckardstein A, Remaley A, Rye KA. HDL in the 21st Century: A Multifunctional Roadmap for Future HDL Research. Circulation. 2021;143(23):2293–309.

